# COVID Faster R-CNN: A Novel Framework to Diagnose Novel Coronavirus Disease (COVID-19) in X-Ray Images

**DOI:** 10.1101/2020.05.14.20101873

**Authors:** Kabid Hassan Shibly, Samrat Kumar Dey, Md. Tahzib-Ul-Islam, Md. Mahbubur Rahman

## Abstract

COVID-19 or novel coronavirus disease, which has already been declared as a worldwide pandemic, at first had an outbreak in a small town of China, named Wuhan. More than two hundred countries around the world have already been affected by this severe virus as it spreads by human interaction. Moreover, the symptoms of novel coronavirus are quite similar to the general flu. Screening of infected patients is considered as a critical step in the fight against COVID-19. Therefore, it is highly relevant to recognize positive cases as early as possible to avoid further spreading of this epidemic. However, there are several methods to detect COVID-19 positive patients, which are typically performed based on respiratory samples and among them one of the critical approach which is treated as radiology imaging or X-Ray imaging. Recent findings from X-Ray imaging techniques suggest that such images contain relevant information about the SARS-CoV-2 virus. In this article, we have introduced a Deep Neural Network (DNN) based Faster Regions with Convolutional Neural Networks (Faster R-CNN) framework to detect COVID-19 patients from chest X-Ray images using available open-source dataset. Our proposed approach provides a classification accuracy of 97.36%, 97.65% of sensitivity, and a precision of 99.28%. Therefore, we believe this proposed method might be of assistance for health professionals to validate their initial assessment towards COVID-19 patients.

## 1. INTRODUCTION

The outbreak of novel coronavirus, which is known as COVID-19, has created an alarming situation all over the world. Coronaviruses are enveloped non-segmented positive-sense RNA viruses belonging to the family *Coronaviridae* and the order *Nidovirales* and broadly distributed in humans and other mammals^**1**^. However, viruses that are responsible for Middle East Respiratory Syndrome (MERS) and Severe Acute Respiratory Syndrome (SARS) also belong to the coronavirus family.^**2, 3**^

The outbreak of COVID-19 started in Wuhan, a town of Eastern China, in December 2019. This virus causes Pneumonia with symptoms such as fever, dry cough, and fatigue. In severe cases, the patient feels difficulty in breathing. Some patients also experience headaches, nausea, or vomiting. It spreads from person to person through droplets of cough or sneeze of an infected person^**4**^. Even an uninfected person touches the droplets and then touches his face, especially eyes, nose, or mouth without washing hands can be infected by this novel coronavirus. As of May 8, 2020, according to the situation report of the World Health Organization (WHO), there are 210 countries affected by the novel coronavirus. On April 25, 2020, it was declared as a pandemic by the World Health Organization (WHO). Reverse Transcription Polymerase Chain Reaction (RT-PCR) is one of the foremost methods of testing coronavirus. This test is performed on respiratory samples, and the results are generated within a few hours to two days. Antibodies are also used to detect COVID-19, where blood samples are used to identify the virus. However, Health professionals use Chest X-Ray scans occasionally to specify lung pathology. In Wuhan, a study was performed on computerized tomography (CT) image reports, and it found that the sensitivity of CT images for the COVID-19 infection rate was about 98% compared to RT-PCR sensitivity of 71%^**5**^. At the early stage of this global pandemic, the Chinese clinical centers had insufficient test kits. Therefore, doctors recommend a diagnosis only based on clinical chest CT results^**6, 7**^.

Even countries like Turkey uses CT images due to the insufficient number of test kits. Some studies state that lab reports and clinical image features are even better for early detection of COVID-19^**8, 9, 10, 11**^. Moreover, health experts also noticed changes in X-Ray images before the symptoms are visible^**12**^. Deep Neural Network approach techniques have had successful application to many problems in recent times, such as skin cancer classification^**13, 14**^, breast cancer detection^**15, 16**^, brain disease classification^**17**^, pneumonia detection in the chest X-Ray^**18**^, and lung segmentation^**19, 20**^. Therefore, precise, accurate, and faster intelligence detection models may help overcome this problem in the rapid rise of this COVID-19 epidemic. In this article, we propose a novel framework to detect COVID-19 infection from X-Ray images using Faster Region Convolutional Neural Network (F R-CNN) deep approach. Based on an available benchmark dataset of COVIDx^**21**^we examined X-Ray images reports of COVID-19 along with the reports of patients with other diseases and normal persons. Also, for feature extraction, we have used the VGG-16 network for building our model.

## 2. RELEVANT WORK

Deep learning is a popular area of research in the field of artificial intelligence. It enables end-to-end modeling to deliver promised results using input data without the need for manual feature extraction. The use of machine learning methods for diagnostics in the medical field has recently gained popularity as a complementary tool for doctors. A molecular diagnosis method of novel coronavirus was proposed^**22**^ by developing two 1-step quantitative real-time reverse-transcription PCR assays for detecting regions of the viral genome. In another exploration, authors have analyzed^**23**^ the Epidemiological and clinical features of novel coronavirus and included the records of all COVID-19 infected patients, which were reported by the Chinese Center for Disease Control and Prevention until January 26, 2020. In recent times, many radiological images have been extensively used to detect COVID-19 confirmed cases. Sethy et al.^**24**^ classified the properties obtained from different models of CNN with the SVM Classifier using X-Ray images. Besides, Wang et al.^**25**^ suggested a Deep model for COVID19 patients recognition and achieved an accuracy of 92.4% in the classification of standard classes, non-COVID, and COVID-19 Pneumonia. In another study, a ResNet50^**26**^ model was proposed by Narin *et al*., and it achieved a COVID-19 detection accuracy of 98%. In terms of COVID-19 patients detection using X-Ray images, the Deep model of Ioannis et al.^**27**^ reached a success rate of 98.75% for two classes and 93.48% for three classes. By comprising multiple CNN models, Hemdan et al.^**28**^ have proposed a COVIDX-Net model that is capable of detecting confirmed cases of COVID-19. A transfer learning-based framework has been advised by Karmany et al.^**29**^ to identify medical diagnoses and treatable diseases using image-based deep learning.

## 3. METHODOLOGY

Convolutional Neural Networks (CNN) is a deep neural network-based learning architecture for processing a massive amount of data. Nowadays, it is widely applied for medical imaging analysis. The CNN is used extensively over other Machine Learning methods because it does not need any manual feature extraction as well as does not require specific segmentation. In Figure 1(A), CNN architecture design is shown, which consists of several blocks of input Layer, Hidden Layer (convolutional layer, pooling layer), fully connected layer, and Output Layer. However, for selecting an anchor, the anchors are divided into two categories (positive and negative) with an intersection-over-union (IoU) overlap ratio between ground-truth box and anchor as a classification index. The IoU overlap ratio is defined as in Equation (1).

**Figure 1:**
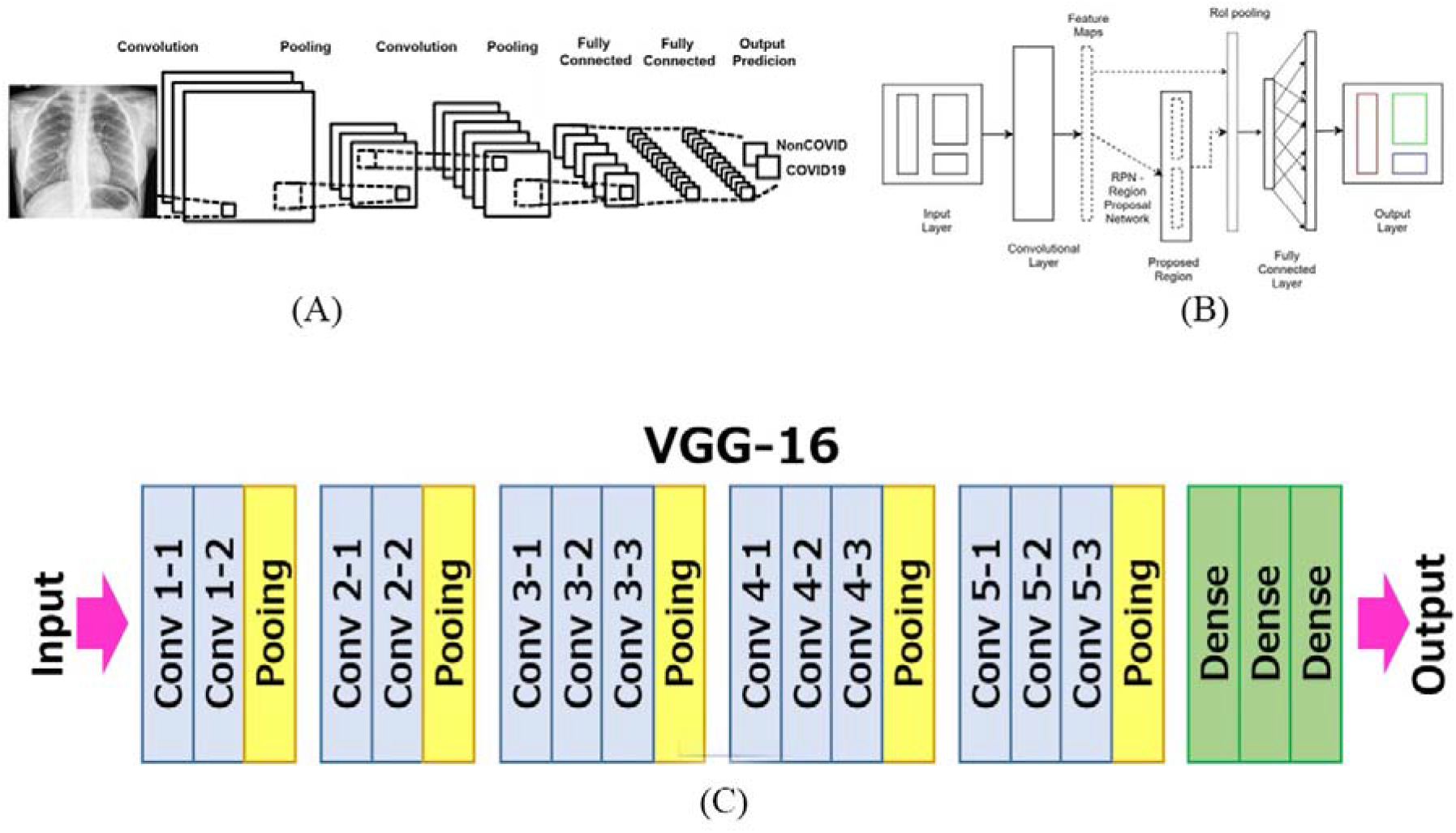
1(A) represents the schematic diagram of a convolutional neural network architecture of XRay image classification; 1(B) showing the pipeline structure of Faster Regional based Convolutional Neural Network (Faster R-CNN), and lastly, 1(C) highlights the basic structure of Convolution, Pooling, and Dense layers of VGG-16 Network Architecture.

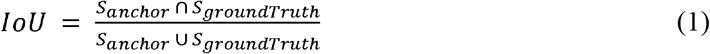

Using VGG-16, we minimize an objective function following the multi-task loss in Faster R-CNN, and the loss function is derived in Equation (2).

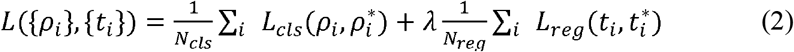

*L_cls_* is the classification loss function, and *L*_reg_ is the regression loss function. *N_cls_* and *N_reg_* are the normalization coefficients of the classification loss function *L_cls_* and the regression loss function *L_reg_* respectively. λ is the weight parameter between *L_cls_* and *L_reg_*. The classification loss function *L_cls_* is the logarithmic loss of two categories (COVID-19 and non-COVID), and it is defined as in the following Equation (3)

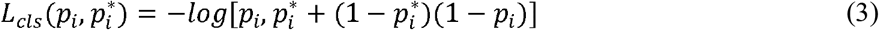

For the regression loss function, it is defined as in Equation (4)

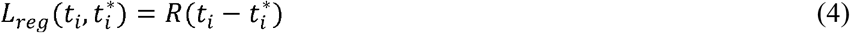

Here, R is defined as a robust loss function in Equation (5)

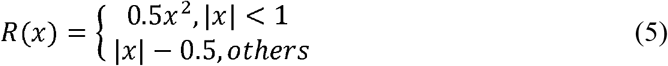

Regarding model development, rather than creating a model from scratch, we built it according to our sample input requirements. We have used similar layers and filters as compared to the original Faster R-CNN architectures and gradually increased the number of filters. In addition, it is essential to consider the Faster R-CNN when analyzing our proposed model and algorithm. Our proposed framework consists of 24 convolutional layers, followed by two fully connected layers and six pooling layers. These layers are typical CNN layers with different filter numbers, sizes, and stride values.

Besides, more modified versions of CNN are available for model development, such as RCNN, Fast R-CNN, and Faster R-CNN. In this exploration, we have used Faster Regional based Convolutional Neural Network (Faster R-CNN). Figure 1(B) is showing the architecture of Faster R-CNN. Furthermore, we used the VGG16 network as the foundation for model classification. We combine this with Faster R-CNN to use this model in a real-time. system. Finally, a deep model with a large number of layers is essential for the extraction of properties of an image in a real-time detection system. For that reason, the model classification structure is capable of grasping and learning small differences. An illustration of the proposed model used in this study is shown in Figure 2.

**Figure 2:**
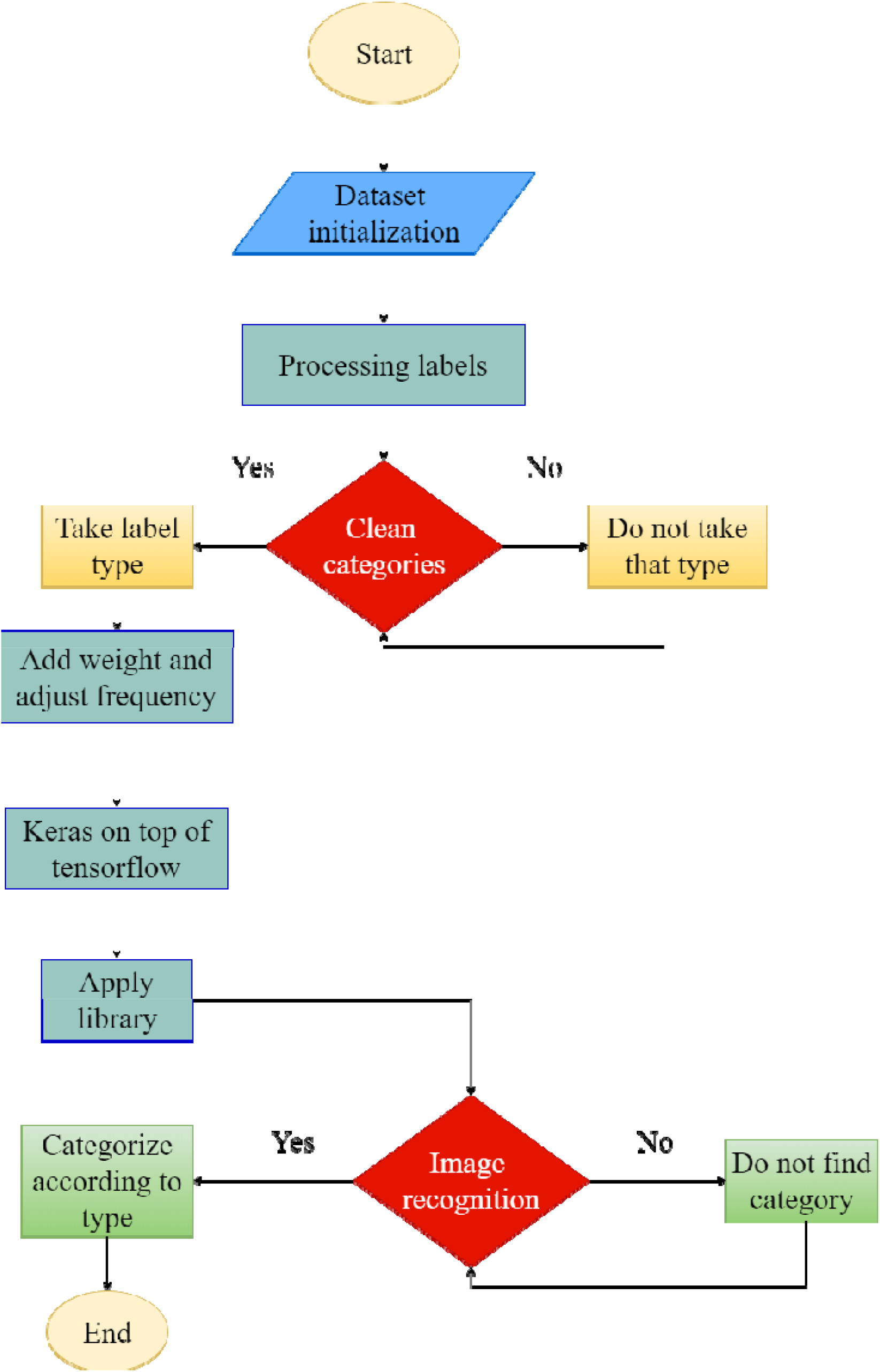
Workflow representation of proposed framework

### 3.1. Data Preparation

For dataset design, we have followed a two-step procedure for data preparation. Initially, we used the X-ray images of COVID-19 patients, which is available as open-source data. Furthermore, we developed a custom dataset to train and evaluate, which comprised a total of 5450 chest radiography images across 2500 patient cases. To prepare the custom dataset for our use in the experiment, we combined and modified two different publicly available datasets: COVID chest X-Ray dataset curated by Dr. Joseph Cohen, a postdoctoral fellow at the University of Montreal^**30**^, and RSNA pneumonia detection challenge dataset from Kaggle^**31**^. In the later phase of this exploration, we moved to a newly available dataset dedicatedly developed for COVID-19 positive case detection using chest X-Ray images named as COVIDx^**22**^. The number of image samples we used for both (custom and COVIDx) the dataset is showing in Table 1. For the custom dataset, we have used 5450 sample images, whereas, in the COVIDx dataset, we developed our dataset with 13800 images.

**Table 1:** Different data representation of multiple datasets used in this research

| Dataset | Type | Normal | non-COVID Pneumonia | COVID19 | Total |
| --- | --- | --- | --- | --- | --- |
| Custom Dataset | Train | 1100 | 3000 | 80 | 4180 |
|  | Test | 300 | 950 | 20 | 1270 |
| COVIDx | Train | 7966 | 5451 | 152 | 13569 |
|  | Test | 100 | 100 | 31 | 231 |

The COVIDx dataset is updated continuously with images shared by researchers from different regions. As of May 7, 2020, there are 183 X-Ray images diagnosed with COVID-19, 8066 patients as normal, and 5551 cases identified as non-COVID Pneumonia. By merging ‘Normal’ and ‘non-COVID Pneumonia’ into a single ‘non-COVID’ class, we designed it as a binary class dataset.

### 3.2. Model Building and Training

The most noticeable fact is that very few patients of COVID-19 associated with X-Ray images, which leads to the scarcity of availability of X-Ray images. For the model building and training, we have used the Googles TensorFlow library and VGG-16 for high-performance numerical computation. Regarding the cross-validation approach, our study used the K-fold cross-validation method (K = 10) with the support of Leave-one-out cross-validation. Algorithm 1 provides insight into K-fold cross-validation working procedures.

#### Algorithm 1: K-fold cross-validation

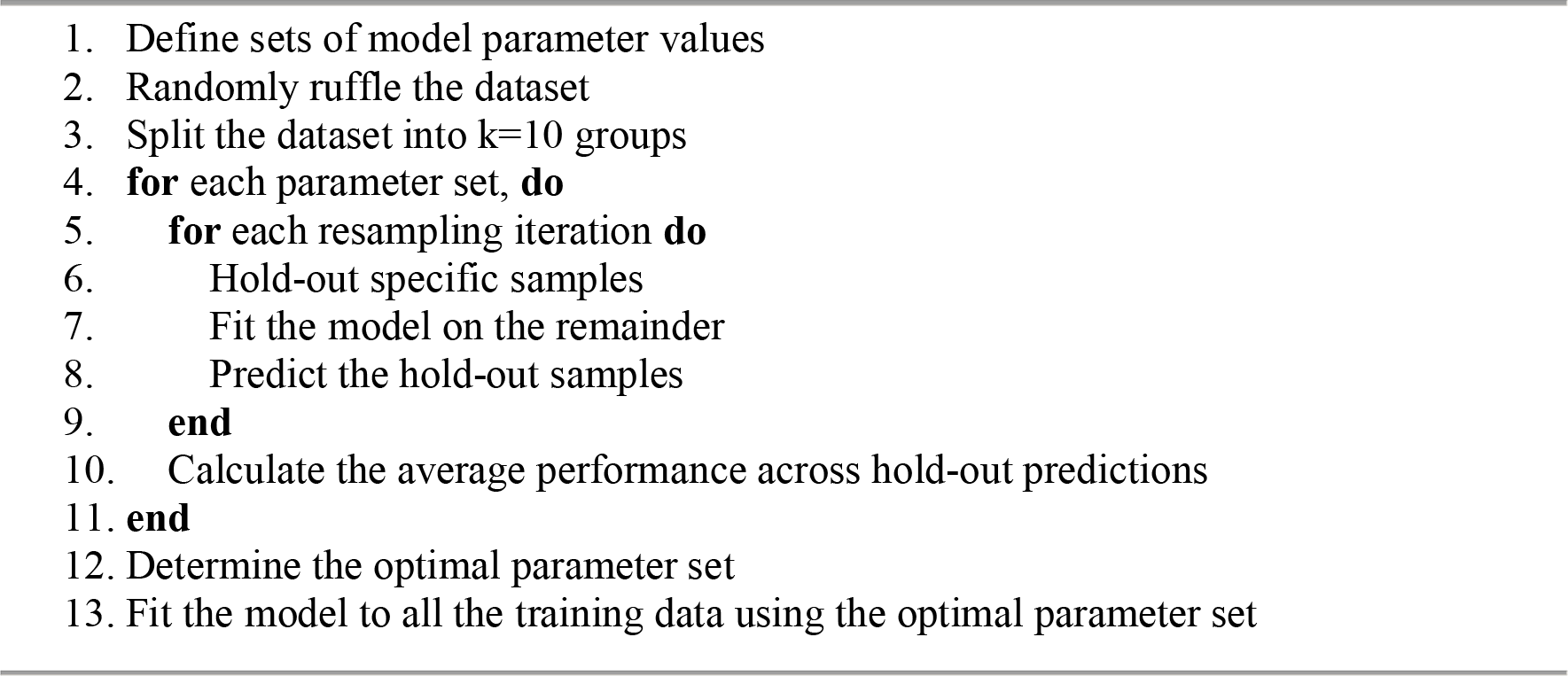
Descriptions of variables in the meta-population model.

Our proposed framework of Faster R-CNN based COVID-19 positive case detection is developed and trained by using the support of Googles Colaboratory. Detection experiment executed on Google’s cloud server’s GPU with 12 GB of RAM and with the support of TensorFlow version 2.1.0. The proposed model was trained with a training learning rate of 2e-5, batch size of 8, and 100 epoch. The complete framework is built and evaluated using the Keras deep learning library with TensorFlow backend^**49**^. All the experiment and data analysis is undergone in the Machine Intelligence Lab (MIL) of Dhaka International University.

## 4. RESULTS

In this section, we discuss the loss observation, followed by the results of model validation. We performed experiments to detect and classify COVID-19 confirmed cases using X-Ray images and train the models in two classes: non-COVID and COVID-19. The model was evaluated using 10-fold cross-validation technique. We have used 90% of X-Ray images for training, and the rest of the 10% are used for testing or validation. Moreover, the loss function is highly essential to understand the excellence of the prediction. From Figure 3., we observe that the training loss and validity loss is decreased gradually after every 100 epoch.

**Figure 3:**
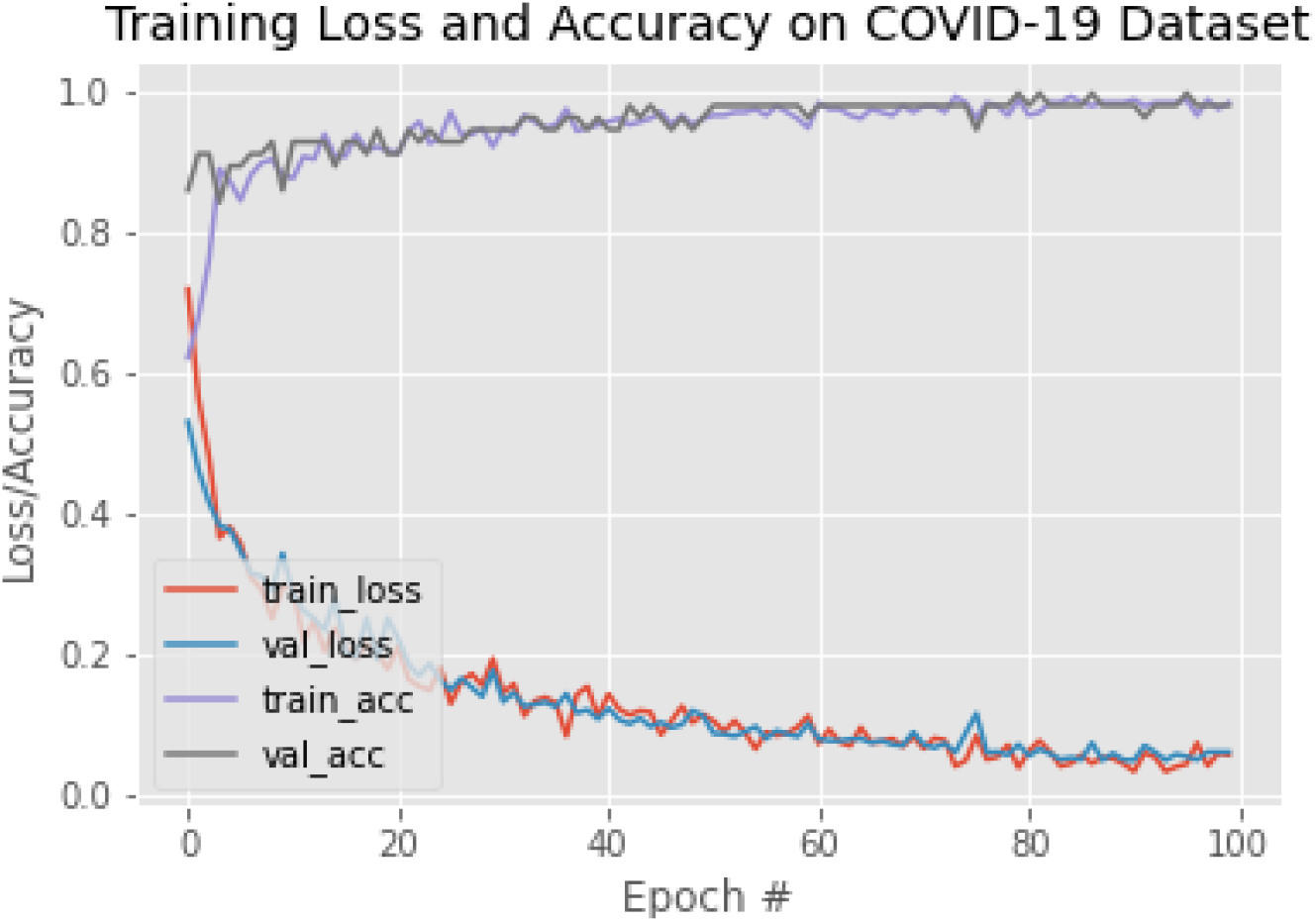
It depicts the loss/accuracy curve, along with the number of Epoch values. It shows that training loss has decreased in every epoch and diverge to a minimum value after 100-epoch. Moreover, validity loss has also reduced like training loss; besides, accuracy on COVID-19 detection has increased after every epoch stage until 100 epoch.

It is also noticeable that both training loss and validation values increased significantly at the primary epoch because of the number of COVID-19 data in that specific class (Figure 3). The effectiveness of the model is achieved through testing, cross-validation, and direct image input testing. In order to evaluate the performance of the model, the complete trained model is validated with the same model dataset using K-Fold cross-validation.

Time comparison is obtained by measuring the time the model takes to complete the detection and classification. Based on the Confusion Matrix four-parameter: True Positive, False Positive, True Negative, and False Negative, our proposed architecture predicted the different samples of Chest X-ray images (Figure 4). The model, however, made incorrect predictions mostly in poor images, and often it predicted the patient with Pneumonia as COVID-19 because they have similarities in image features.

**Figure 4:**
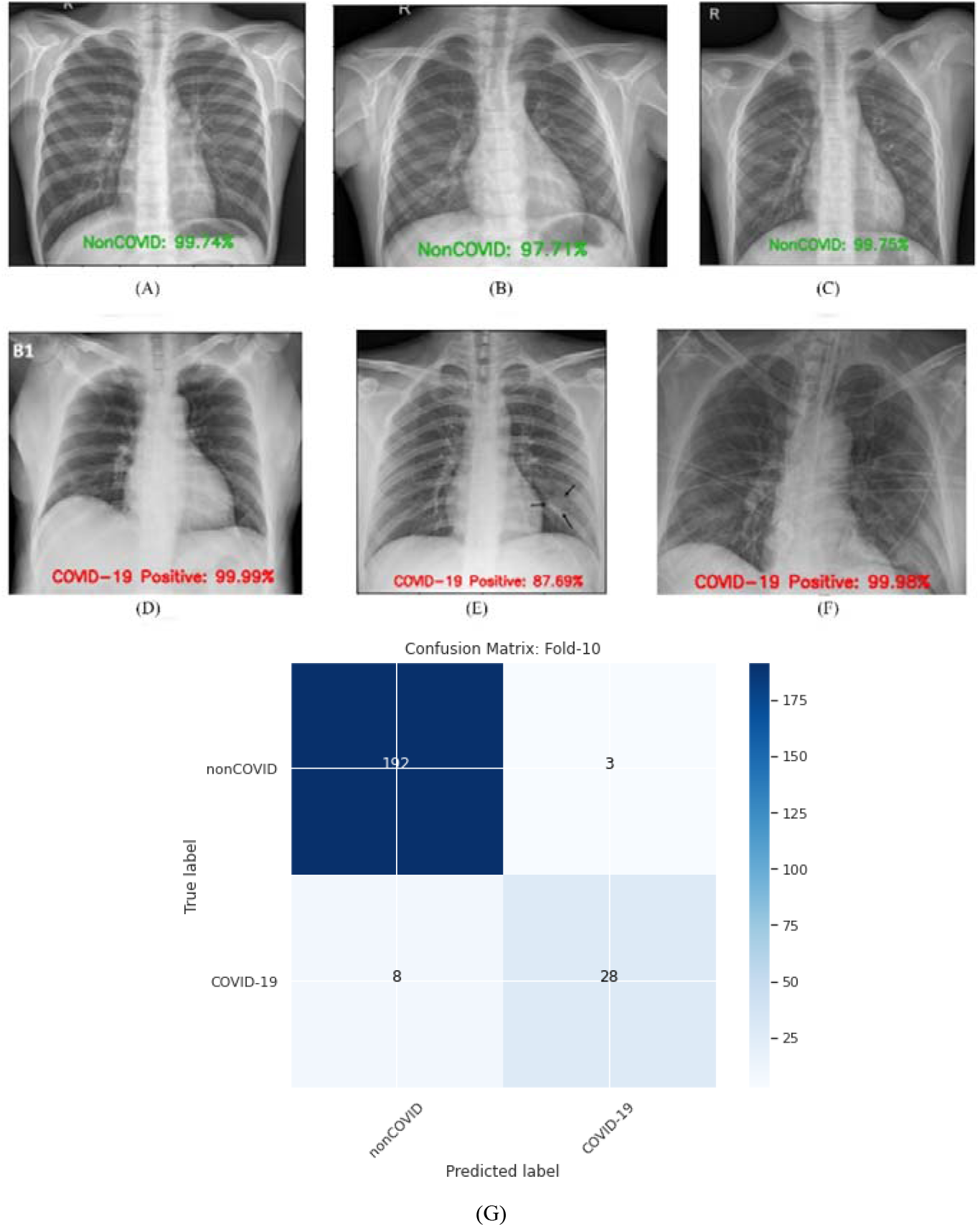
Illustration of different sample images are shown here. Figure 4 (A, B, D, E, and F) shows that the model predicts the sample as non-COVID, whereas the actual class is also a non-COVID. However, only 4 (C) predict the sample as non-COVID, whereas the actual class is COVID-19. 4(G) depicts the generated confusion matrix based on 10-fold cross-validation method.

Performance metrics of the proposed model and 10-fold cross-validation results of different metrics and their average results are tabulated in Table 2. It is observable that the developed model provided an average accuracy of 97.36%, and it obtained an average value of sensitivity, specificity, and F1-score of 97.65%, 95.48%, and 98.46%, respectively. It also provides a precision value of more than 99.00%. We also compare our proposed method with other similar deep learning approaches, and a comparison table has been designed to understand the impact of our framework over other studies. From Table 3, it is evident that Deep CNN ResNet-50^**28**^ based approach produces slightly higher (∼0.64%) detection accuracy in comparison with our proposed method.

**Table 2:** 10 fold cross-validation approach and results of different performance metrics

| Number of Folds | Sensitivity % | Specificity % | Precision % | F1-score % | Accuracy % |
| --- | --- | --- | --- | --- | --- |
| Fold-1 | 98.50 | 96.77 | 99.49 | 98.99 | 98.27 |
| Fold-2 | 99.50 | 100.00 | 100.00 | 99.75 | 99.57 |
| Fold-3 | 97.00 | 93.55 | 98.98 | 97.98 | 96.54 |
| Fold-4 | 99.50 | 93.55 | 99.00 | 99.25 | 98.70 |
| Fold-5 | 98.50 | 100.00 | 100.00 | 99.24 | 98.70 |
| Fold-6 | 97.50 | 93.55 | 98.98 | 98.24 | 96.97 |
| Fold-7 | 97.00 | 96.77 | 99.49 | 98.23 | 96.97 |
| Fold-8 | 96.50 | 90.32 | 98.47 | 97.47 | 95.67 |
| Fold-9 | 96.50 | 100.00 | 100.00 | 98.22 | 96.97 |
| Fold-10` | 96.00 | 90.32 | 98.46 | 97.22 | 95.24 |
| **Average** | **97.65** | **95.48** | **99.29** | **98.46** | **97.36** |

**Table 3:**
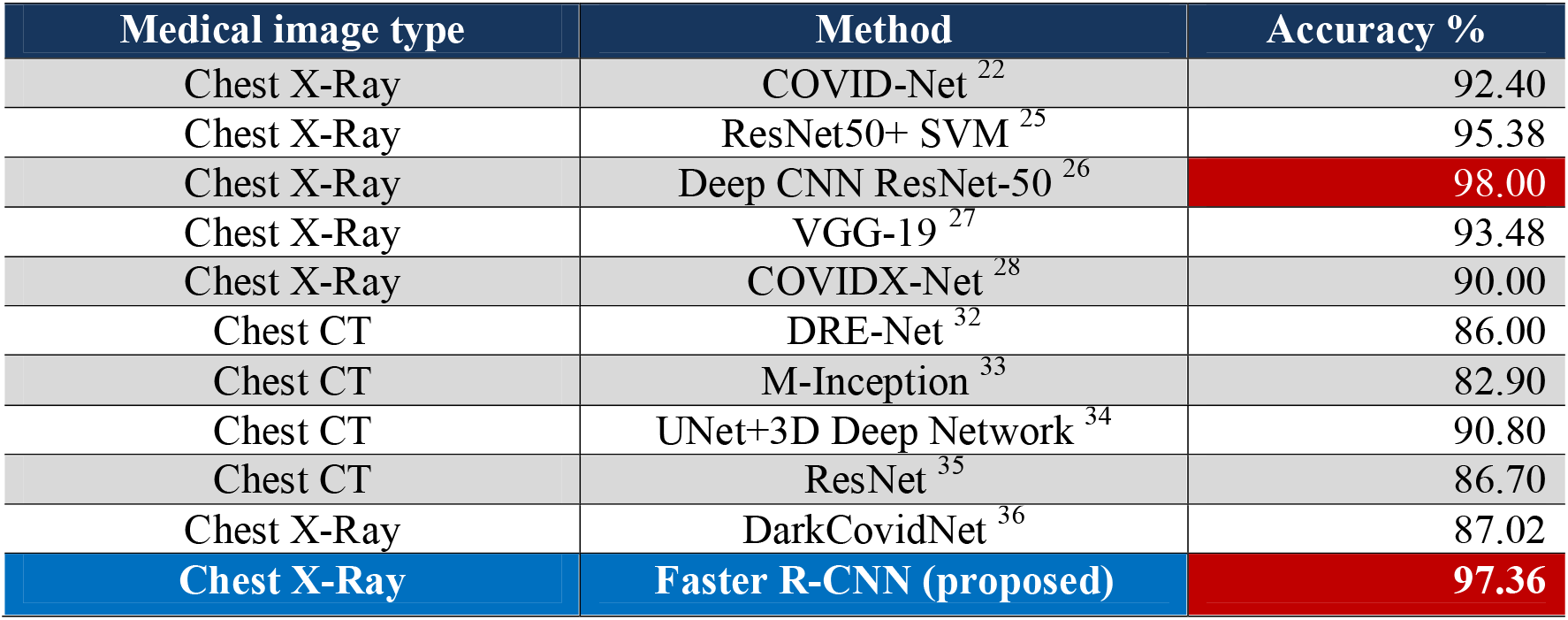
Comparison of the proposed framework with other deep learning approaches

## 5. CONCLUSION

In this study, we have proposed a deep learning model to detect COVID-19 cases from Chest X-Ray images. This automated system can perform binary classification without manual feature extraction with an accuracy of 97.36%. Moreover, this model is also capable of testing with a larger dataset and work with real-time systems. Furthermore, it can be helpful in areas where the test kit is not sufficient. Therefore, for the initial assessment of COVID-19 patients, this tool can act as a fruitful medium of diagnosis under the supervision of radiologists and doctors. At this point, we are working to make our model more vigorous so that it can detect both CT and X-Ray images.

## Data Availability

All data generated or analysed during this study are included in this published article. Wang, L. and Wong, A., 2020. COVID-Net: A tailored deep convolutional neural network design for detection of COVID-19 cases from chest radiography images. arXiv preprint arXiv:2003.09871.

## Author Contribution

SKD and KHS had the idea for and designed the study and had full access to all the data in the study and take responsibility for the data and accuracy of the model generation. SKD, KHS, and MR contributed to the writing of the article. MR and TUI contributed to the critical revision of the report. All the data preparation and models developed by SKD and KHS. All authors contributed to data acquisition, data analysis, result validation, and reviewed and approved the final version.

## Funding

None

## Declaration of Interests

All authors declare no competing interest

## Ethical Approval

Not required

## Notes

### Competing Interest Statement

The authors have declared no competing interest.

